# Aiding Oral Squamous Cell Carcinoma diagnosis using Deep learning ConvMixer network

**DOI:** 10.1101/2022.08.18.22278971

**Authors:** Nguyen Quoc Toan

## Abstract

In recent years, Oral squamous cell carcinoma (OSCC) has become one of the world’s most prevalent cancers, and it is becoming more prevalent in many populations. The high incidence rate, late diagnosis, and inadequate treatment planning continue to be major concerns. Despite the enhancement in the applications of deep learning algorithms for the medical field, late diagnosis, and approaches toward precision medicine for OSCC patients remain a challenge. Due to a lack of datasets and trained models with low computational costs, the diagnosis, an important cornerstone, is still done manually by oncologists. Although Convolutional neural networks (CNNs) have become the dominant architecture for vision tasks for plenty of years, recent experiments show that Transformer-based models, most noticeably the Vision Transformer (ViT), may out-compete them in some settings. Therefore, in this research, a method called ConvMixer, which combines great features from CNNs and patches based on ViT was applied to an original very small dataset of only 1224 images in total for 2 classes, Normal epithelium of the oral cavity (Normal) and OSCC, 696 slides for 400x magnification and 528 slides for 100x magnification. However, the proposed models with small parameters and data augmentation performed magnificently, with 400x magnification (Accuracy: 99.81% - F1score: 99.87%) and 100x magnification (Accuracy: 99.62% - F1score: 99.77%).

## 1. INTRODUCTION

Oral squamous cell carcinoma (OSCC) is a hazardous disease. Its etiology is multifactorial, with tobacco smoking and alcohol consumption being the most significant risk factors. Predominantly affects older people, mostly men, beginning at the age of 40 and peaking at the age of 60. Nowadays, it is becoming more widespread, particularly among younger patients. The question of whether young and old patients with OSCC different prognoses have is still debatable. The tongue and the floor of the mouth are two of the main locations. Surgical treatment has traditionally been the preferred primary cornerstone therapy for it. Furthermore, given the aggressive nature of OSCC and the fact that many patients are diagnosed with locoregionally advanced disease, multi-modality therapy is used to treat them with concurrent chemo-radiotherapy is required. Despite the treatment options mentioned above, the high incidence rate and suboptimal treatment outcome remain a major concern. Early detection is critical for better prognosis, treatment, and survival. This is vital for improving cancer management. Therefore, deep machine learning techniques have been touted for improving early detection and, as a result, lowering cancer-specific mortality and morbidity. Automated image analysis has the potential to aid pathologists and clinicians in the early detection of OSCC and in making informed cancer management decisions.

Natural image classification has developed significantly in accuracy in recent years, thanks to the increasing use of convolutional neural networks (CNNs) and vision transformer (ViT) techniques. And meanwhile, this technology has been used to detect mitosis in histological sections of breast cancer, detect skin cancer, evaluate mammograms, and classify cytology in peripheral blood. However, successful use of CNNs for image classification typically necessitates a sufficient amount of high-quality image data and high-quality annotation, which can be difficult to obtain due to the high cost of obtaining medical expert labels. This is especially true when there is no underlying technical gold standard and human examiners are required to provide ground truth labels for network training and evaluation, such as in the case of OSCC cytomorphologic examination.

Hence, I represent a small dataset of 1224 expert-annotated images taken with a Leica ICC50 HD microscope, from 230 patients at Ayursun-dra Healthcare Pvt. Ltd Center and Dr. Bhubaneswar Borooah Cancer Institute (a government of India-approved regional cancer center) with normal oral cavity epithelium and oral squamous cell carcinoma. In terms of the number of diagnoses, patients, and cell images included, it is, to my knowledge, the largest image data set of Oral epitheliums available in the literature. Therefore, it is a resource that can be used for educational purposes as well as future approaches to automated image based OSCC classification. I separated the dataset to train 2 CNNs-ViT-based (ConvMixer) classifiers for microscope images of OSCC and the normal ones, the first model using 100x magnification and the second one with 400x magnification. I tested and found that they perform admirably, achieving high levels of accuracy for clinically relevant classes while minimizing parameters and computational costs.

## 2. RELATED WORK

OSCC classification is a binary classification task that necessitates the use of machine learning or deep learning techniques to learn how to label images with a class label, in short, 0 or 1. An easy-to-understand explanation is classifying slides as ‘cancerous’ (OSCC) or ‘not cancerous’ (Normal) which is taken from human. Some efficient methods were applied to conduct the dataset [2] as a classification task. Here is a summary of the related work, along with comments and shortcomings.

**Table 1:**
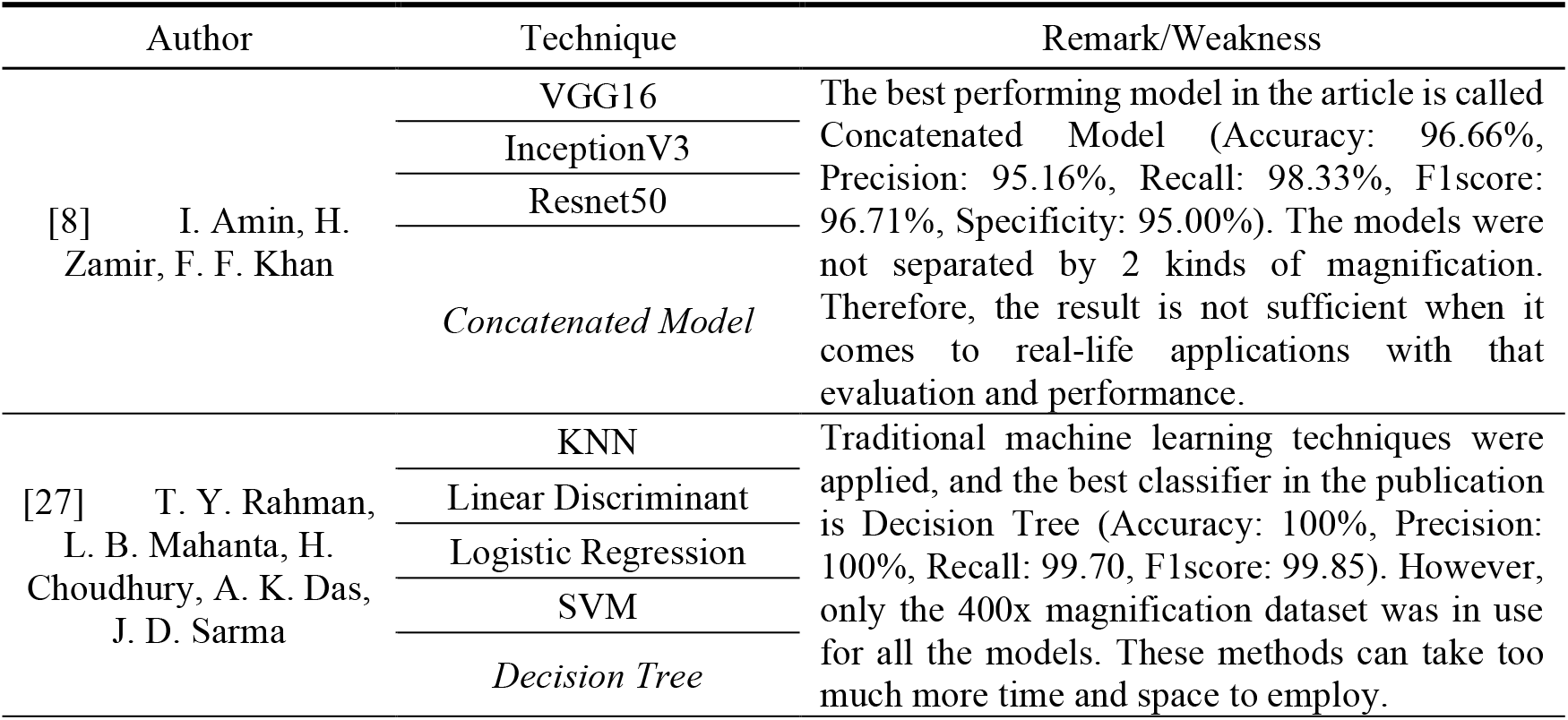
Summary of the related work

## 3. METHODOLOGY

### 3.1 Overview of Conxmixer network

Convmixer network is the magnificent combination of CNNs and Vision transformer (ViT). CNNs make use of pixel arrays, whereas ViT divides images into visual embeddings. The visual transformer then divides an image into fixed-size tokens and passes positional embedding to the transformer encoder as an input. It essentially represents an image as a set of words or word embeddings (patches). It divides the image into patches or tokens rather than using pixels like a CNN. This begged the question of whether its superiority was due to the transformer architecture or the use of patches rather than pixel arrays. That is the reason why the ConvMixer comes out, proves the latter point, and innovates a simple but super effective network.

Patches are fed into Convmixer, which separates spatial and channel mixing, and ensures that all patches are the same size and resolution across the network. However, it employs convolutions to achieve the mixing steps. It also employs batch normalizations rather than layer normalizations. Despite using standard convolutions, it outperforms its competitors as well as basic CNN models by a wide margin [1]. A patch embedding layer is at the core of the ConvMixer model, which is nothing more than a convolution layer with c input channels and h output channels, kernel size, and stride equal to the patch size. This is followed by an activation function and batch normalization after activations. It is illustrated in a detailed way with Figure (6). This is the model’s first component.

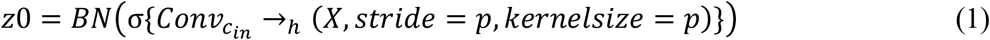

The main ConvMixer layer, which is repeated depth times, is the second part of the model. This layer is composed of residual blocks with a depthwise convolution. A residual block is simply a block that adds the output of a previous layer to the output of a subsequent layer. In this case, the inputs are concatenated to the depthwise convolution output layer. Following this output is an activation block, a Pointwise convolution, and another activation block.

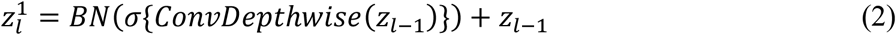

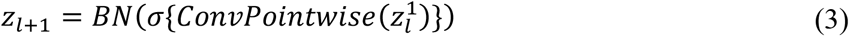

The third component of the model is a global pooling layer that generates a feature vector of size h, which we can then pass to a Softmax classifier or any other head depending on the task.

### 3.2 GELU

GELU, or Gaussian Error Linear Unit, is the activation function used throughout the model. Unlike RELU, the GELU activation function weighs inputs based on their magnitude rather than gating them based on their sign.

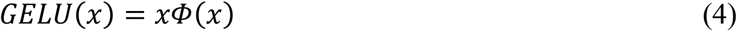

### 3.3 Depthwise Separable

The article [25] was the first to introduce depthwise separable convolution. Depthwise separable convolution is a type of factorized convolution that combines standard convolution (SC) with a pointwise convolution (PWC) and a depthwise convolution (DWC). It has fewer parameters than ‘standard’ convolutional layers, making it less susceptible to overfitting. With fewer parameters, it also necessitates fewer operations to compute, making it cheaper and faster. Figure (1) shows how standard convolution, depthwise convolution, and pointwise convolution work. In standard convolution, each input channel must perform a convolution with a specific kernel, with the result being the sum of all convolution results.

Depthwise convolution is the first step in depthwise separable convolution, with convolution performed separately for each input channel. It is a type of convolution that employs only one convolutional filter for each input channel. Standard convolution, on the other hand, is performed over multiple input channels, and the filter is as deep as the input, allowing us to freely mix various channels to generate the output. These channels are kept apart by depthwise convolution. It also does not change the depth of the image by adding new channels. It is primarily used to blend the spatial dimensions of an image.

A type of convolution in which a 1×1 kernel iterates every pixel or point in the image is known as Pointwise convolution. The depth of this kernel, like the depth of a normal convolution, is equal to the number of input channels. Pointwise convolution can be used to increase the number of image channels (filters). Its primary function is to blend data across patches.

The number of weights required for standard convolution, as shown in Figure (1), with an input feature map of size M x M x N and a kernel size of K x K x N x P is calculated by the equation (5). And the number of operations that corresponds to it is equation (6). The total number of weights in depthwise separable convolution is equation (7), as well as the total number of operations in equation (8).

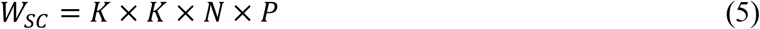

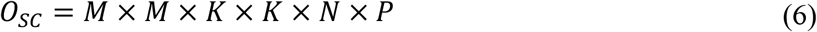

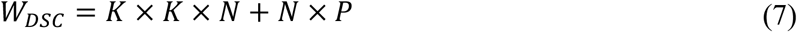

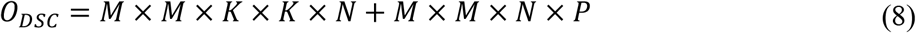

**Figure 1:**
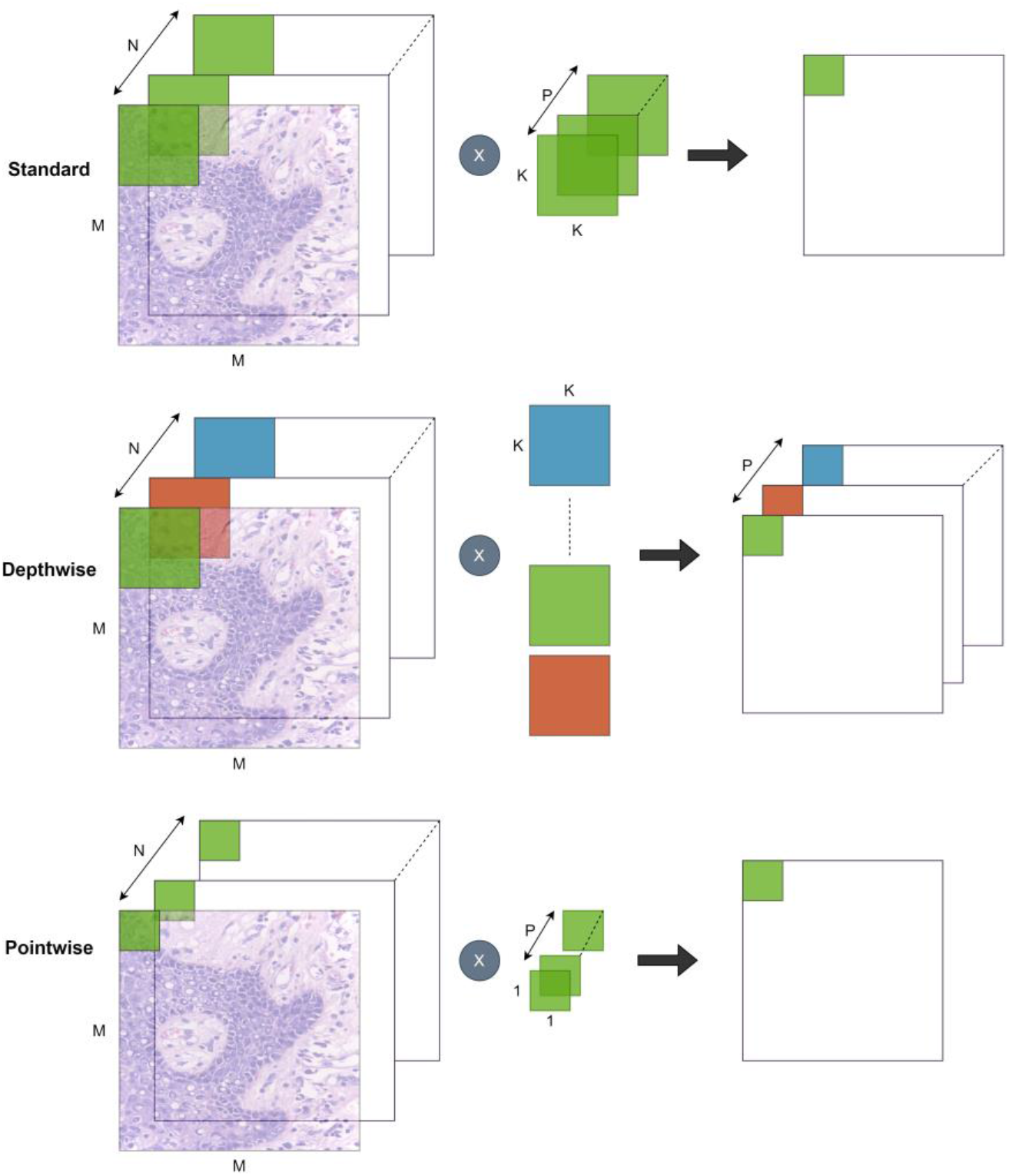
Illustration of Standard, Depthwise, and Pointwise convolution

Therefore, operation and reduction factors on weights are calculated in equation (9) (10):

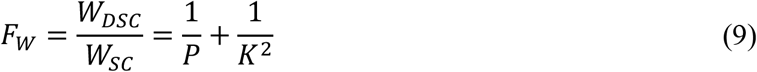

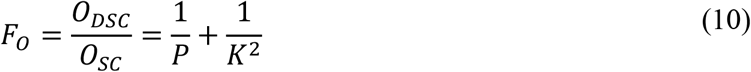

## 4. DATASET

The dataset was obtained from two sources: Ayursundra Healthcare Pvt. Ltd. and Dr. B Borooah Cancer Research Institute, both of which have been authorized as regional cancer centers by Indian government. The images were viewed and captured using a Leica ICC50 HD microscope (size 2048×1536), dataset information is from reference [2]. Following the data augmentation steps (Flip, Rotate, Blur, Noise), the images were resized to 256×256, along with thousands of augmented data samples, to achieve a higher level of model performance.

**Table 2:** Summary of datasets for training models

| Dataset | Class | Total image |
| --- | --- | --- |
| 100x magnification | Normal | 1335 |
|  | OSCC | 6585 |
| 400x magnification | Normal | 3015 |
|  | OSCC | 7425 |

**Figure 2:**
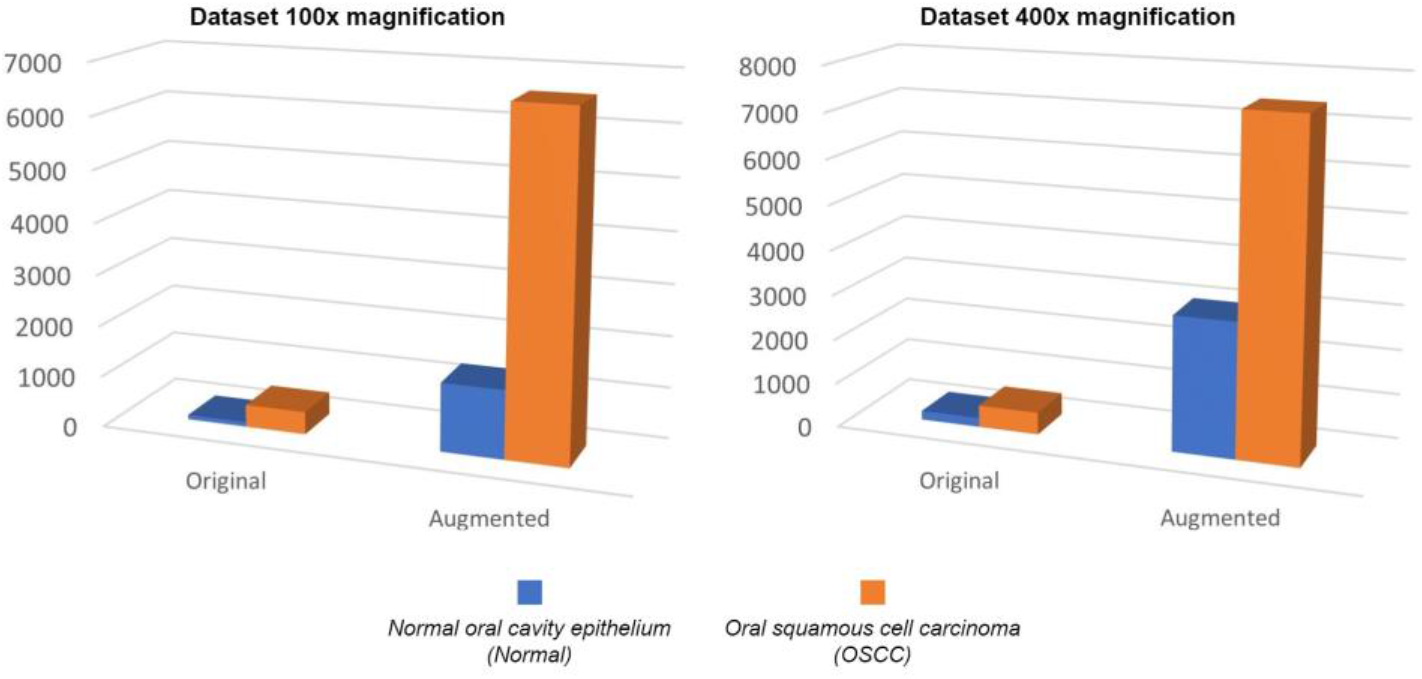
Comparison between original datasets and augmented datasets for 100x and 400x magnification

**Figure 3:**
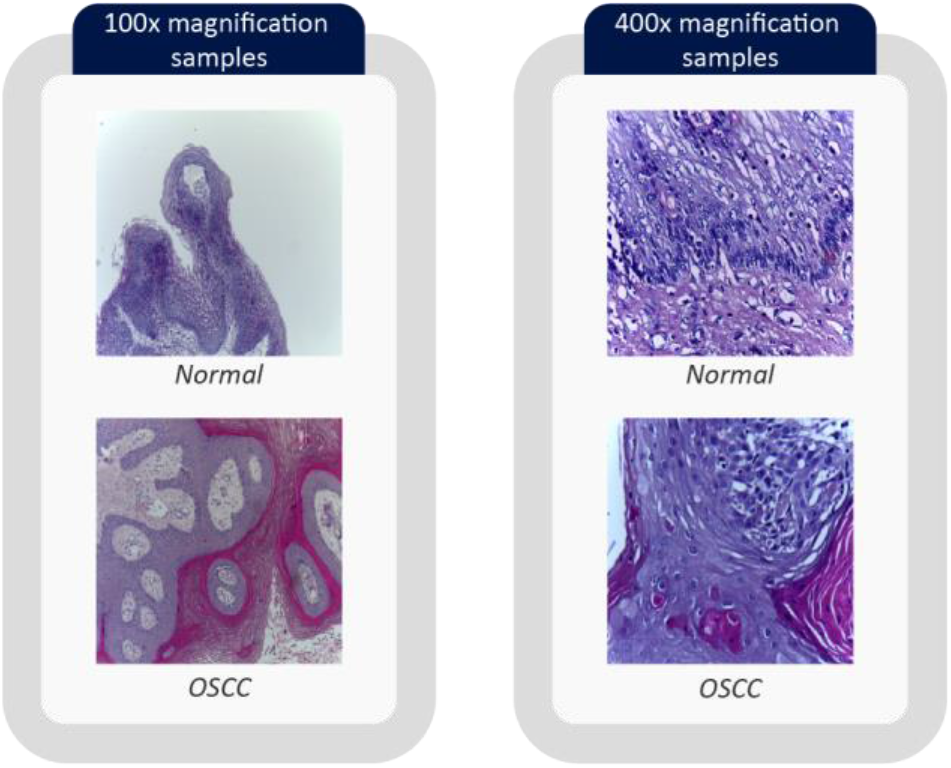
Slide samples

## 5. EXPERIMENT

### 5.1 Flowchart

The proposed research processes are depicted in the flowchart below:

**Figure 4:**
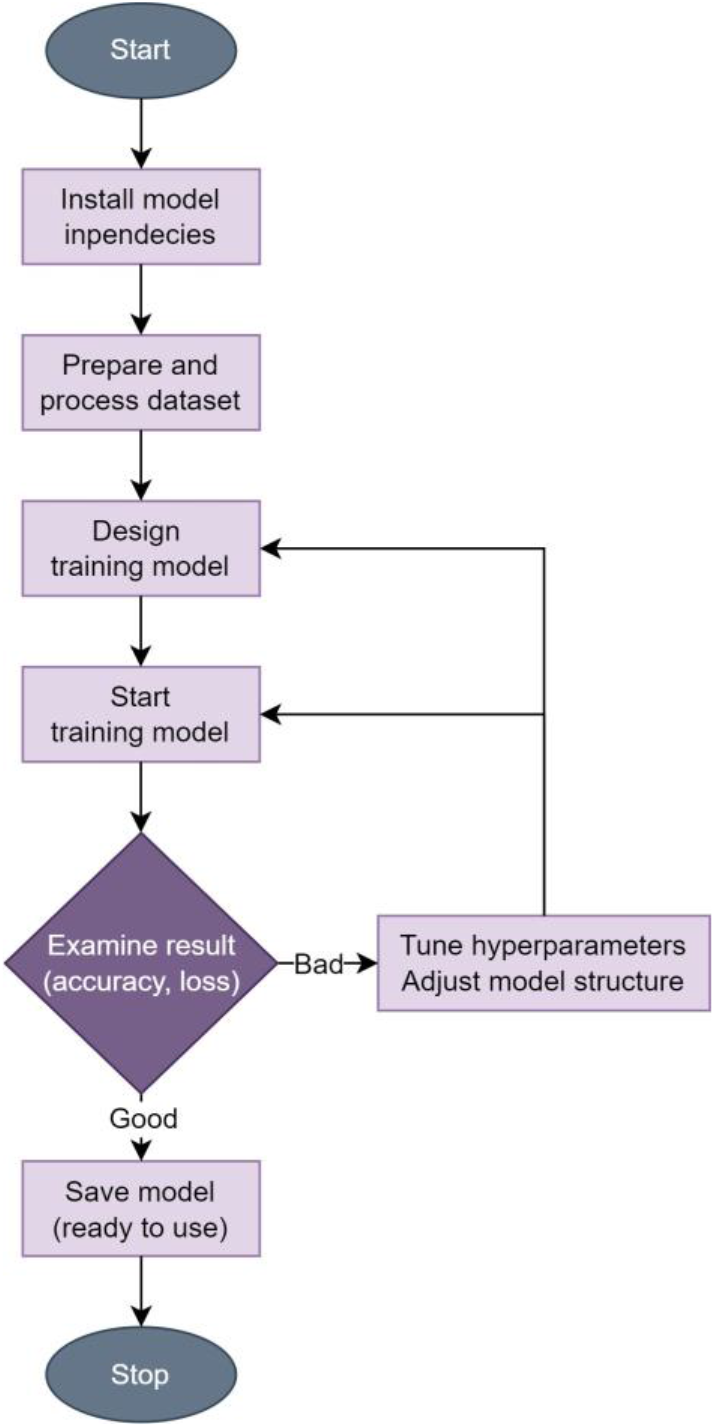
Flowchart of the proposed research

### 5.2 Training

The training was carried out on 2 GPUs - NVIDIA GeForce RTX 2080 Ti (11GB-GDDR6) by the algorithm:

**Figure 5:**
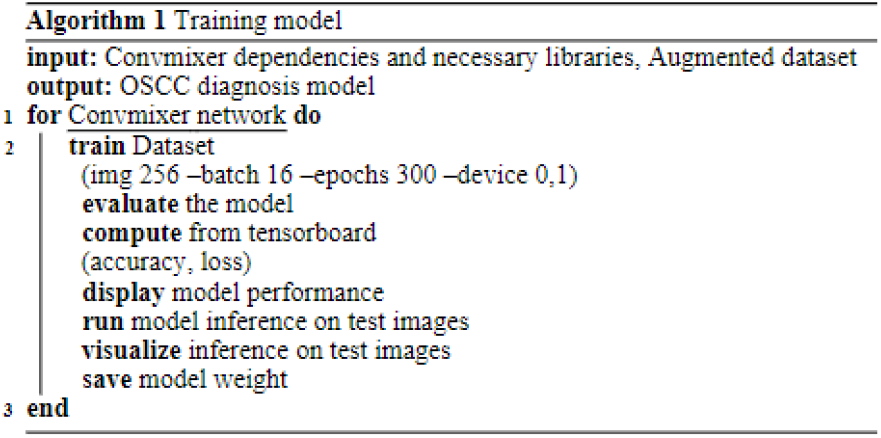
The proposed algorithm

### 5.3 The proposed Conmixer network

Figure (6) is an overview of how the proposed method works with a 256×256 input size. First and foremost, it embeds the image into patches, a process is known as ‘Patch Embedding’. The workflow is followed from top to bottom, with the final output being Normal or OSCC.

**Figure 6:**
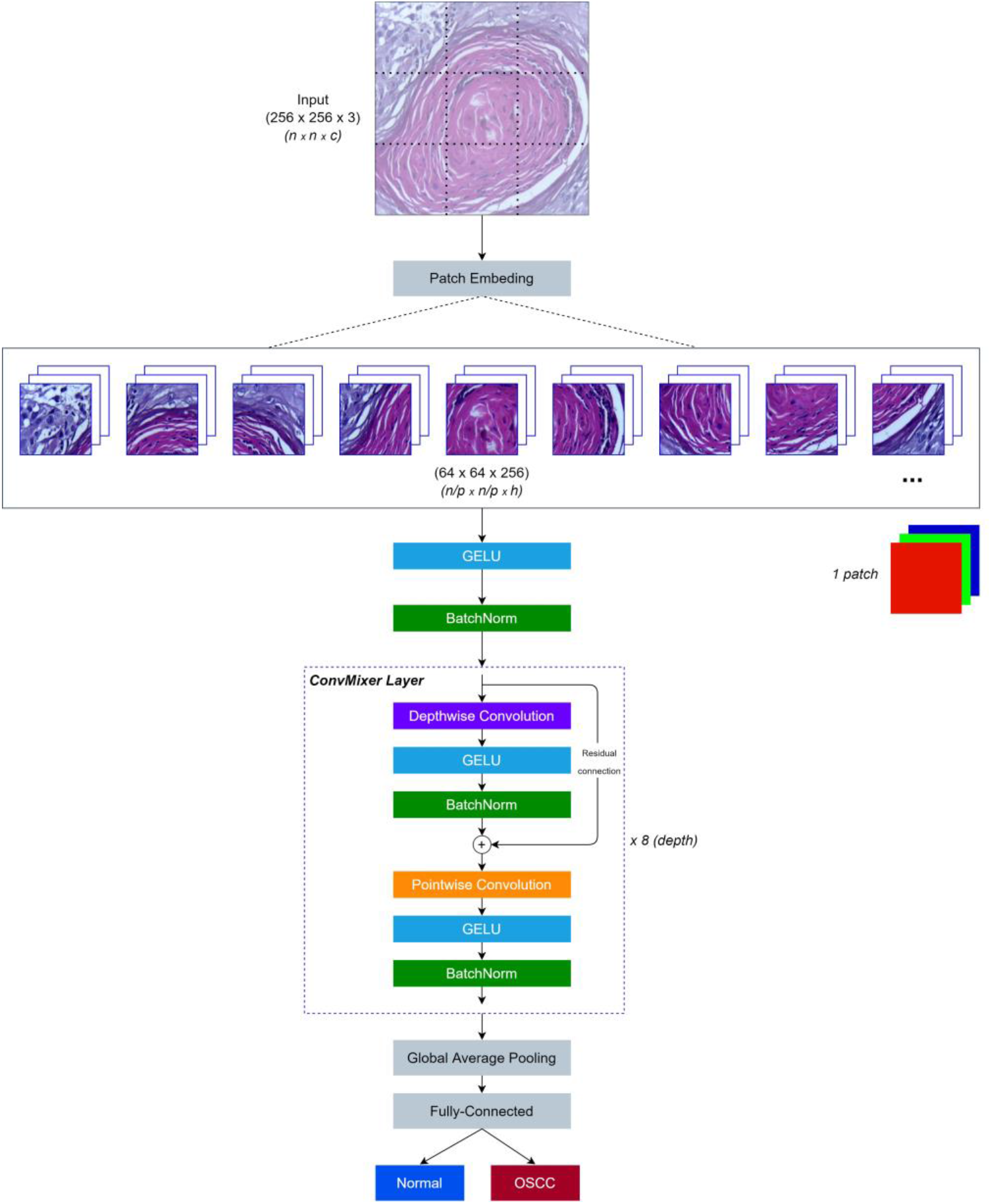
Illustration of the proposed Convmixer network

**Figure 7:**
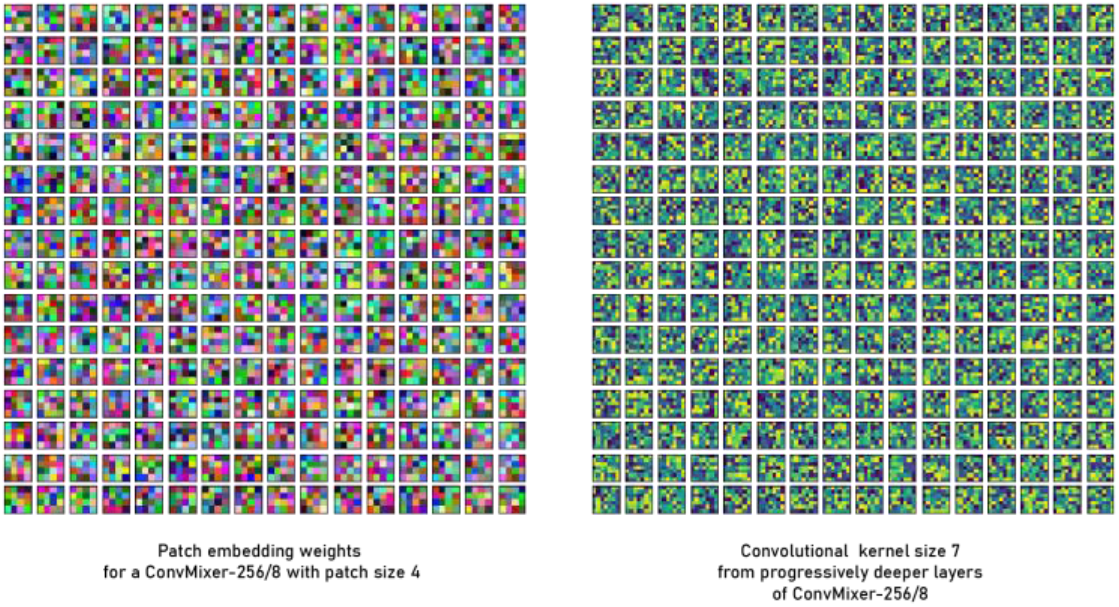
Patch embedding weights - convolutional kernel (Internal Convmixer network)

## 6. RESULT

### 6.1 Evaluation metrics

Accuracy, Precision, Recall, Specificity, and F1score were used to evaluate the models. True Positive (TP) in OSCC classification is the OSCC was correctly classified, False Positive (FP) predicted OSCC and the actual class was not. False Negative (FN) occurs when the predicted class is Normal, and the actual class is OSCC. True Negative (TN) means that the predicted class was Normal, and the actual class was Normal.

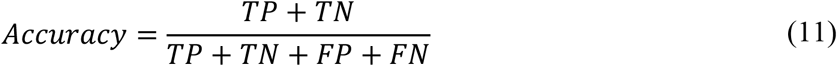

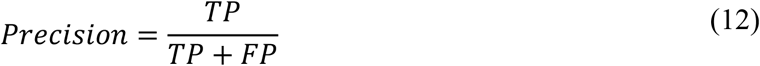

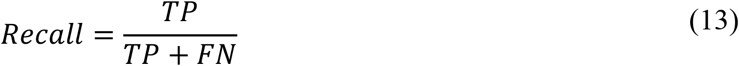

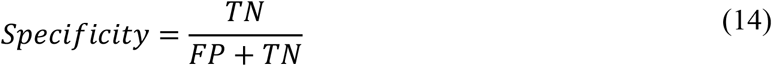

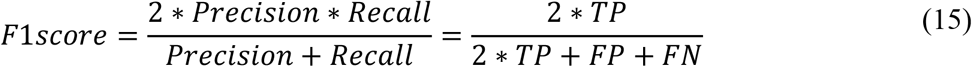

### 6.2 Model results

To evaluate, 793 slides (658 OSCC and 135 Normal) for 100x magnification and 1043 (742 OSCC and 301 Normal) were used to test the two proposed models which are 10% of each dataset.

**Figure 8:**
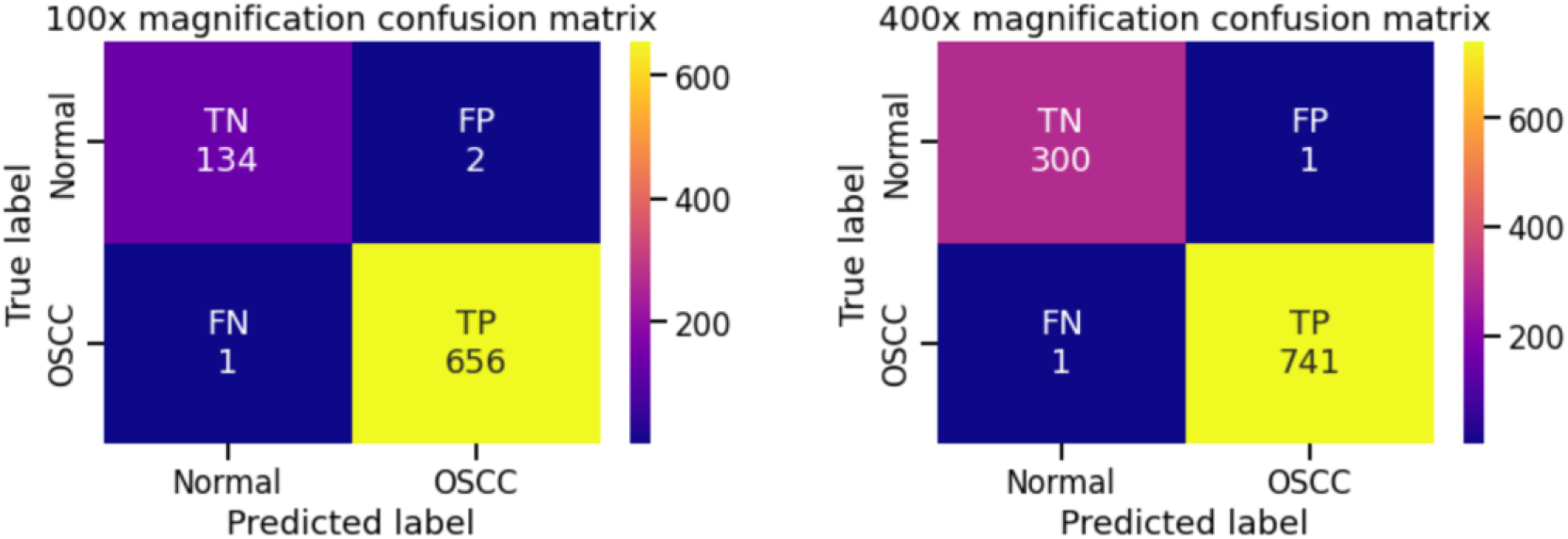
Confusion matrix of the proposed models on test datasets

**Table 3:** Models results in summary

| Metric \ Model | 100x magnification | 400x magnification |
| --- | --- | --- |
| **Input size** | 256x256 | 256x256 |
| **Patch Size** | 4 | 4 |
| **Kernel Size** | 7 | 7 |
| **Depth** | 8 | 8 |
| **Params ($\times 10^3$)** | 659 | 659 |
| **Accuracy (%)** | 99.62 | 99.81 |
| **Recall (%)** | 99.85 | 99.87 |
| **Specificity (%)** | 98.53 | 99.67 |
| **Precision (%)** | 99.70 | 99.87 |
| **F1score (%)** | 99.77 | 99.87 |
| **Loss** | 0.039 | 0.038 |

**Figure 9:**
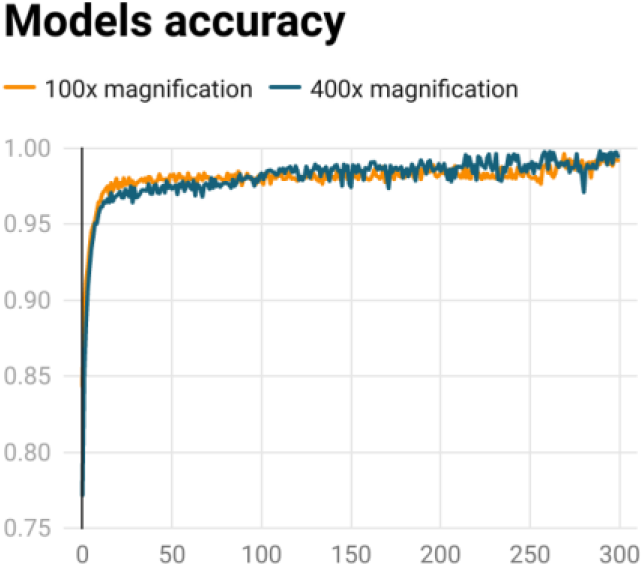
The training process for models’ accuracy

**Figure 10:**
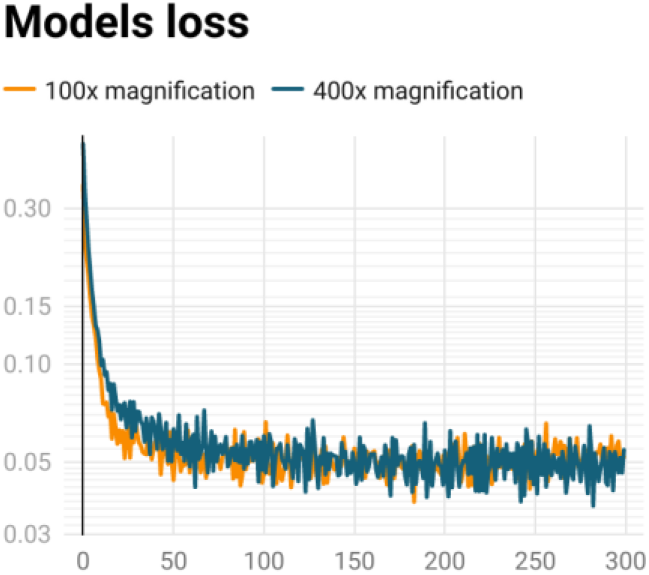
The training process for models’ loss

## 6. CONCLUSION

In this paper, the purpose is to attempt to distinguish between normal and OSCC microscopic images of oral squamous cell carcinoma from histologist slides. For classification, the Convmixer network was used, and the results proved to be very satisfactory and almost perfect with evaluated metrics is being achieved with a very simple network structure, resulting in low computational costs. The method will act as an effective platform for the development of computer-aided diagnostic tools to aid clinicians/pathologists in the rapid differentiation of tumorous lesions from normal ones. The efficiency of the network structure and applied methods were proved by the results with only 300 epochs and achieved exceptional results with 400x magnification (Accuracy: 99.81% - F1score: 99.87%) and 100x magnification (Accuracy: 99.62% - F1score: 99.77%). If the datasets are more plentiful, they will perform better than this proposed research. Furthermore, Convmixer should be a popular deep learning network for cell classification tasks with small datasets.

Yet, a small amount of test data may not completely represent the data collected in the real world, and training data may not have adequate coverage of the distribution. This study’s OSCC dataset was both small and unbalanced. It is usable and acceptable, but it is not ideal for translating to real-world applications. The models only can be applied as an assistance tool. Future enhancement of this study is to improve this in the future by generating large datasets with matched classes. Correspondingly, combining the structures of other networks which are based on CNNs and ViT should be applied for enhancing the model on larger datasets.

## Data Availability

Total 1224 images are there in this repository. Images are divided into two sets in two different resolutions. First set composed of 89 histopathological images with the normal epithelium of the oral cavity and 439 images of Oral Squamous Cell Carcinoma (OSCC) in 100x magnification. The second set consists of 201 images with the normal epithelium of the oral cavity and 495 histopathological images of OSCC in 400x magnifications. The images were captured using a Leica ICC50 HD microscope from H&E stained tissue slides collected, prepared and cataloged by medical experts from 230 patients. 

https://data.mendeley.com/datasets/ftmp4cvtmb/1

## ACKNOWLEDGEMENT

I would like to send my heartfelt gratitude to HAIL (Hongik University - Artificial Intelligence Laboratory, Seoul, Republic of Korea) which is advised by Professor Seongwon Cho for supporting me to employ this research.

